# SARS-CoV-2 detection using digital PCR for COVID-19 diagnosis, treatment monitoring and criteria for discharge

**DOI:** 10.1101/2020.03.24.20042689

**Authors:** Renfei Lu, Jian Wang, Min Li, Yaqi Wang, Jia Dong, Weihua Cai

## Abstract

**Background:** SARS-CoV-2 nucleic acid detection by RT-PCR is one of the criteria approved by China FDA for diagnosis of COVID-19. However, inaccurate test results (for example, high false negative rate and some false positive rate) were reported in both China and US CDC using RT-PCR method. Inaccurate results are caused by inadequate detection sensitivity of RT-PCR, low viral load in some patients, difficulty to collect samples from COVID-19 patients, insufficient sample loading during RT-PCR tests, and RNA degradation during sample handling process. False negative detection could subject patients to multiple tests before diagnosis can be made, which burdens health care system. Delayed diagnosis could cause infected patients to miss the best treatment time window. False negative detection could also lead to prematurely releasing infected patients who still carry residual SARS-CoV-2 virus. In this case, these patients could infect many others. A high sensitivity RNA detection method to resolve the existing issues of RT-PCR is in need for more accurate COVID-19 diagnosis.

**Methods:** Digital PCR (dPCR) instrument DropX-2000 and assay kits were used to detect SARS-CoV-2 from 108 clinical specimens from 36 patients including pharyngeal swab, stool and blood from different days during hospitalization. Double-blinded experiment data of 108 clinical specimens by dPCR methods were compared with results from officially approved RT-PCR assay. A total of 109 samples including 108 clinical specimens and 1 negative control sample were tested in this study. All of 109 samples, 26 were from 21patients reported as positive by officially approved clinical RT-PCR detection in local CDC and then hospitalized in Nantong Third Hospital. Among the 109 samples, dPCR detected 30 positive samples on ORFA1ab gene, 47 samples with N gene positive, and 30 samples with double positive on ORFA1ab and N genes.

**Results:** The lower limit of detection of the optimize dPCR is at least 10-fold lower than that of RT-PCR. The overall accuracy of dPCR for clinical detection is 96.3%. 4 out 4 of (100 %) negative pharyngeal swab samples checked by RT-PCR were positive judged by dPCR based on the follow-up investigation. 2 of 2 samples in the RT-PCR grey area (Ct value > 37) were confirmed by dPCR with positive results. 1 patient being tested positive by RT-PCR was confirmed to be negative by dPCR. The dPCR results show clear viral loading decrease in 12 patients as treatment proceed, which can be a useful tool for monitoring COVID-19 treatment.

**Conclusions:** Digital PCR shows improved lower limit of detection, sensitivity and accuracy, enabling COVID-19 detection with less false negative and false positive results comparing with RT-PCR, especially for the tests with low viral load specimens. We showed evidences that dPCR is powerful in detecting asymptomatic patients and suspected patients. Digital PCR is capable of checking the negative results caused by insufficient sample loading by quantifying internal reference gene from human RNA in the PCR reactions. Multi-channel fluorescence dPCR system (FAM/HEX/CY5/ROX) is able to detect more target genes in a single multiplex assay, providing quantitative count of viral load in specimens, which is a powerful tool for monitoring COVID-19 treatment.

## 1. Introduction

Coronavirus disease 2019 (COVID-19)^1^ is now becoming a global public health problem, as the definition of “the first pandemic in history that could be controlled” nominated by WHO^2^. Severe acute respiratory syndrome coronavirus 2 (SARS-CoV-2)^3^, the pathogen of COVID-19, was first isolated and sequenced in early January 2020^4^. One-step reverse-transcription real-time PCR^5^ is recommended by the Chinese Center for Disease Control and Prevention (CDC) as the “gold standard” for diagnosis of COVID-19^6^. However, this commonly used method showed relatively lower sensitivity (about 30%-50%) than expected. That could be partly due to low viral load in the pharyngeal of some patients, the inappropriate transport and storage of samples, and relatively low detection limit of RT-PCR. Patients with symptoms of COVID-19 but false negative detection of SARS-CoV-2 by RT-PCR may be treated as if they were suffering regular flu or pneumonia, which results in high risk of viral transmission and high mortality^7^. One of the requirements of discharging convalescent, “two consecutive days’ negative detection for SARS-CoV-2 by RT-PCR”, may also lead to potential risk of viral transmission^6^. Therefore, a more sensitive detection method is required for accurate SARS-CoV-2 diagnosis.

The concept of digital PCR was conceived and described in 1992^8^, and first published as “digital PCR” in 1999^9^. The absolute quantification results come from Poisson statistics after limited dilution and endpoint PCR^10,11^. This method is also more robust against PCR inhibitors existing broadly in clinical samples^12^. The superior precision of digital PCR could be used for the detection of small fold change of copy number variation or gene expression^13^. Digital PCR was also applied for rare mutation detection in cancer diagnostics^14^, because the abundance of rare mutation in a partition is relatively high and easier to detect^15^ than in bulk.

Here, we demonstrated the application of dPCR assay showing higher sensitivity by one order of magnitude than RT-PCR. The dPCR assay can be used as a complementary method to the RT-PCR detection method. Based on the results of this optimized dPCR system, we showed that the overall accuracy of the dPCR for clinical SARS-CoV-2 detection is 96.3%.

### Ethics statement

The Ethics Committee of the Nantong Third Hospital Affiliated to Nantong University approved this study. Existing samples collected during standard diagnostic tests were tested and analyzed retrospectively by dPCR. No extra burden was posed to patients.

## 2. Materials and Methods

### 2.1. Clinical samples and RNA extraction

The samples were obtained from clinical patients with fever, coughing, or lung inflammation confirmed by CT images at Nantong Third Hospital Affiliated to Nantong University and local CDC. Pharyngeal swabs were soaked in 1000 μl PBS buffer. RNA from the pharyngeal swabs was extracted using Liferiver Bio-Tech automatic nucleic acid extractor (Model: EX3600/2400) following manufacturer’s instruction.

### 2.2. Primers and probes

The primers and probes targeted the ORF1ab and N of SARS-CoV-2 according to Chinese CDC. Target 1 (ORF1ab), forward: 5’-CCCTGTGGGTTTTACACTTAA-3’, reverse: 5’-ACGATTGTGCATCAGCTGA-3’, probe: 5’-FAM-CCGTCTGCGGTATGTGGAAAGGTTATGG-BHQ1-3’; Target 2 (N), forward: 5’-GGGGAACTTCTCCTGCTAGAAT-3’, reverse: 5’-CAGACATTTTGCTCTCAAGCTG-3’, probe: 5’-HEX-TTGCTGCTGCTTGACAGATT-TAMRA-3’. Internal references gene (RPP30), forward: 5’-AGT GCA TGC TTA TCT CTG ACA G-3’, reverse: 5’-GCA GGG CTA TAG ACA AGT TCA-3’, probe: 5’-Cy5-TTT CCT GTG AAG GCG ATT GAC CGA-BHQ-3’.

### 2.3. Workflow

For dPCR workflow, all the procedures follow the manufacturer’s instructions of RainSure DropX-2000 Droplet Digital PCR System using RainSure Novel Coronavirus (SARS-CoV-2) Nucleic Acid Detection Kit. Briefly, 25μlof reaction mix is required for each reaction on the DropX-2000 platform. Each 25μlis comprised of 10 μl of SARS-CoV-2 one-step RT digital PCR master mix, 1 µl of enzyme mix, 14 μl RNA extracted from patient samples. Following the instrument touch screen’s prompt, microfluidic droplet generation and detection cartridges were placed on a cartridge loading stage. 70 μl of droplet generation oil and 25 μl of reaction mix were loaded into an oil well and a sample well, respectively. After the reagent loading process, a gasket with filters was mounted onto the wells of the reagent loaded cartridge. The instrument retrieved the cartridge loading stage and started droplet generation process automatically followed by a thermal cycling protocol: step 1, 49°C for 20 min (reverse transcription); step 2, 97 ° C for 12 min (DNA polymerase activation); step 3, 40 cycles of 95.3°C for 20 sec (denaturation) and 52°C for 1 min (annealing); step 4, 20°C (cooling) for infinite hold. The cartridges were then transferred and loaded onto the DScanner-2000 for multi-channel fluorescence detection of droplets. A single multiplex assay measures the concentration of 3 different target genes, ORF1ab, N gene and RPP30 respectively.

For RT-PCR workflow, the primers and probes are from Liferiver Bio-Tech. A25-μlreaction was set up containing 5 μl of RNA, 19 μl of reaction buffer provided with the one step RT-PCR system and 1µl enzyme mix. Thermal cycling was performed at 45°C for 10 min for reverse transcription followed by 95°C for 3 min and then 45 cycles of 95 °C for 15 sec, 58 °C for 30 sec in SLAN 96P real time PCR system.

### 2.4 Data analysis

Analysis of the dPCR data was performed with analysis software GeneCount V1.60b0318 (RainSure Scientific). Concentrations of the target RNA sequences, along with their Poisson-based 95% confidence intervals were also provided by the software. Fluorescence channels of FAM, HEX and Cy5 were scanned to detect ORF1ab gene, N gene and RPP30 gene respectively. The positive populations for each target gene were identified using positive and negative controls with single primer–probe sets for each fluorescence channel. The concentration reported by GeneCount has the unit of copies of template per microliter of the final 1× dPCR reaction, which was also reported and used in all the subsequent analysis.

## 3. Results

### 3.1. Comparison of the lower limit of detection between dPCR and the standard RT-PCR

Lower limit of detection (LLoD) of RT-PCR and dPCR was compared using serial dilution of clinical specimen. The starting clinical specimen showed Ct value of 35 in RT-PCR. The specimen was diluted using virus storage solution. Each dilution was 5 fold. A total of 7 dilutions (8 samples S1-S8 including the starting stock) were tested by both RT-PCR and dPCR assays. As shown in **Figure *1*** and **Table 2** RT-PCR failed to detect S3, while dPCR was able to detect S3 and S4. dPCR showed negative results for S5 through S8. dPCR assay showed at least 10 times lower LLoD than RT-PCR assay. However. LLoQ (lower limit of quantification) was estimated to be larger than the viral concentration in S3.

**Figure 1.**
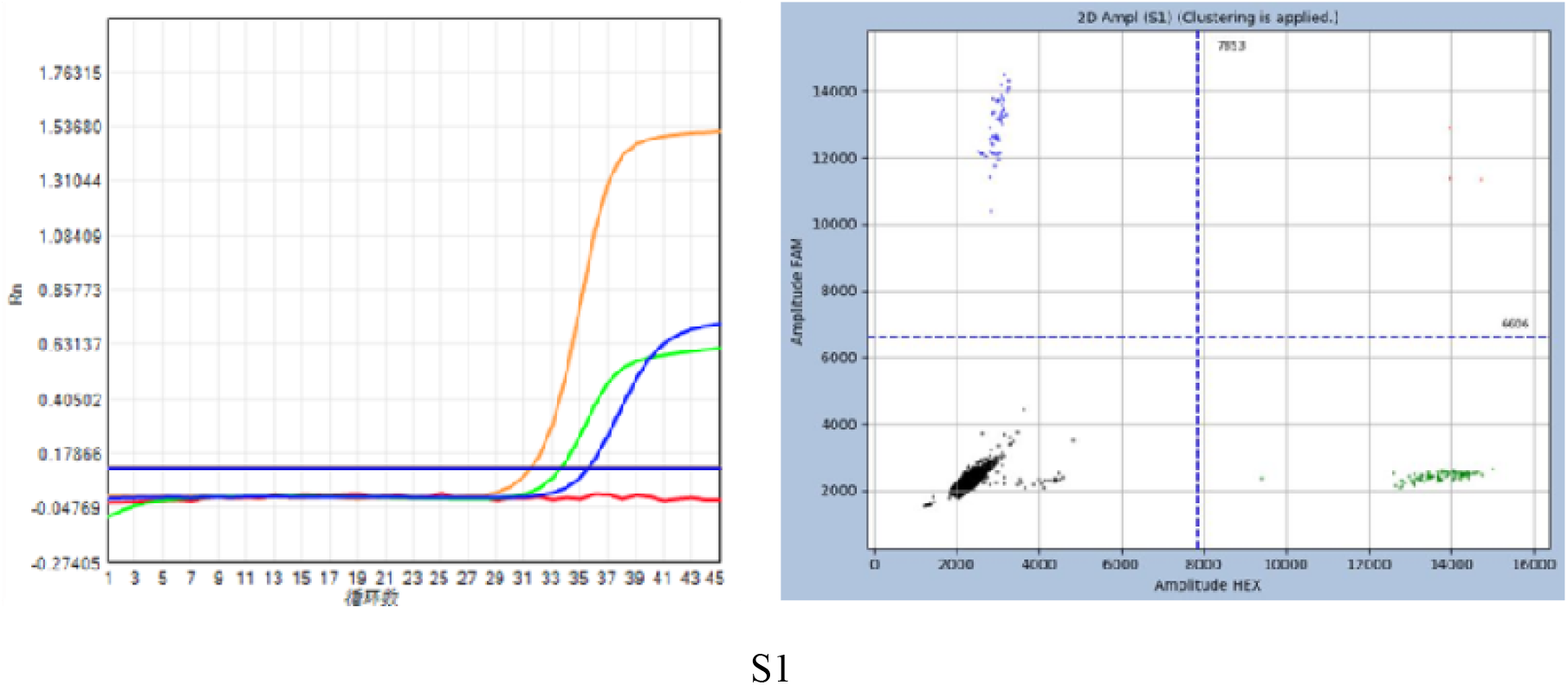
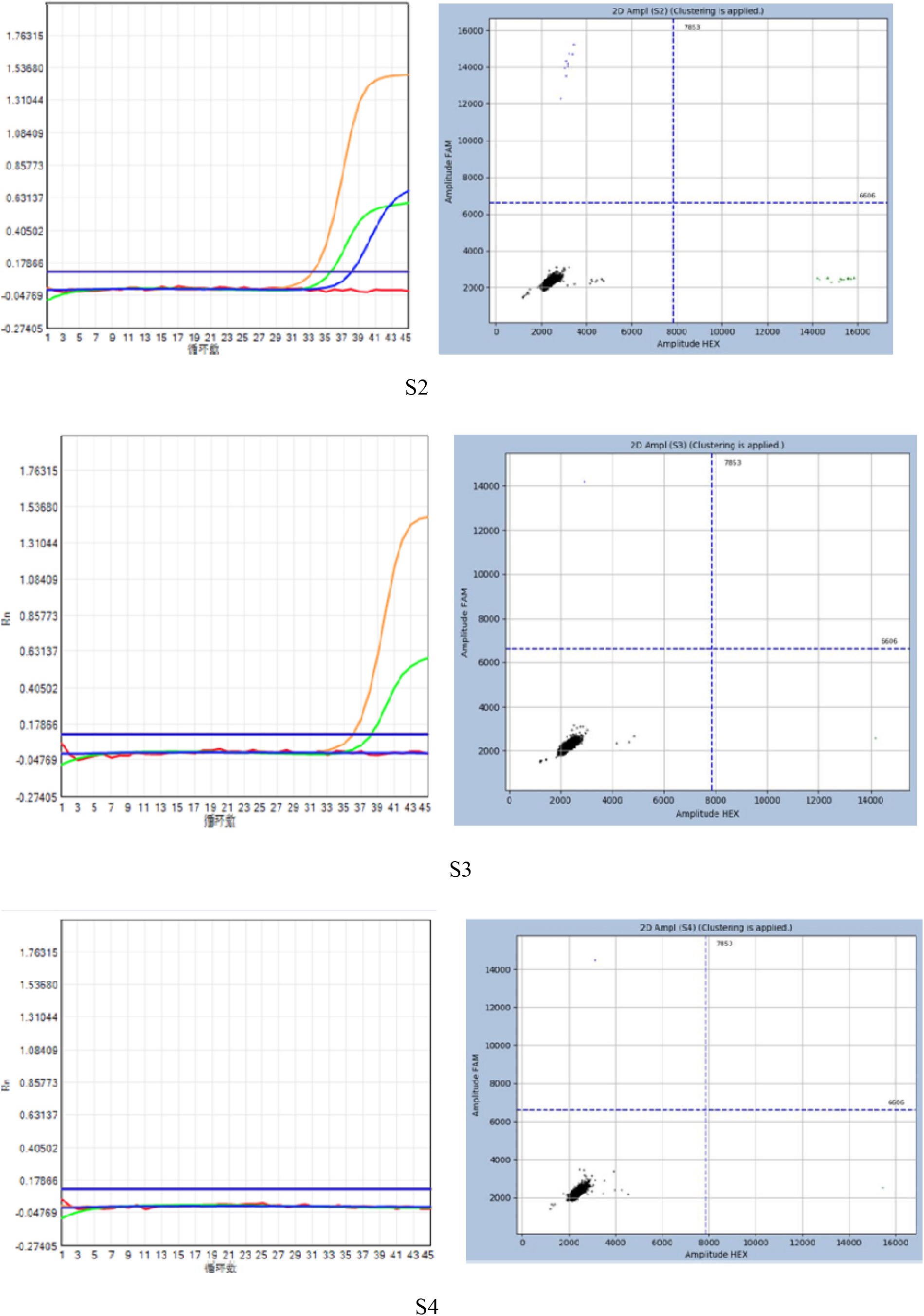
Graphs and data of lower limit of detection (LLoD) of RT-PCR and dPCR using series dilution of clinical specimen. The clinical specimen showed RT-PCR Ct value at 35. The specimen was diluted using virus storing solution 5 for each dilution. Each dilution is 5 fold. Total of 7 dilutions (8 concentrations) were tested by both RT-PCR and dPCR assays. Orange color is internal reference, green is N gene, blue is ORF1ab for both RT-PCR and dPCR. S1 is original specimen (left RT-PCR, right dPCR). S2 is S1 specimen 5X dilution (left RT-PCR, right dPCR). S3 is S2 specimen 5X dilution (left RT-PCR, right dPCR). S4 is S3 5X dilution (left RT-PCR, right dPCR). S5-S8 are 5X series dilutions from S4 specimen. The dPCR assay showed at least 10 fold lower LLoD than RT-PCR assay. RT-PCR failed to detect at S3 dilution, dPCR was able to detect S3 and S4 dilution. However. LLoQ (lower limit of quantification) is above S3 concentration.

**Figure 2.**
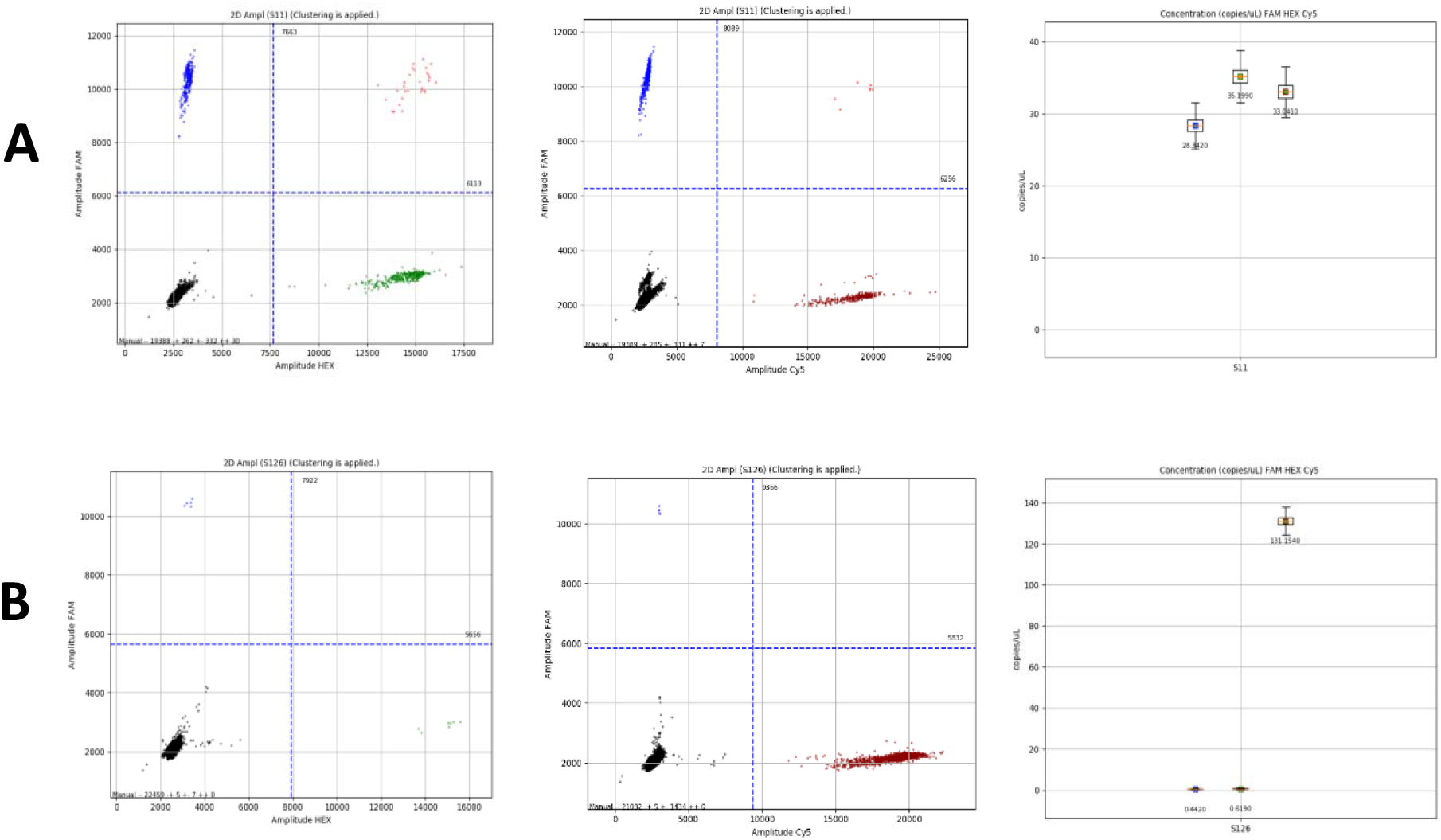
dPCR results of SAR-CoV-2 tests from patient 33. 2A is test result from Patient 33 on Jan. 27^th^, 2020. 2B is test results from patient 33 on Feb 5^th^, 2020. Left is FAM(ORF1ab) Y axis, HEX (N) X axis; middle is FAM (ORF1ab) Y axis, Cy5 (internal reference) X axis; right is the concentration call of ORF1ab, N and internal reference genes.

**Table 1.**
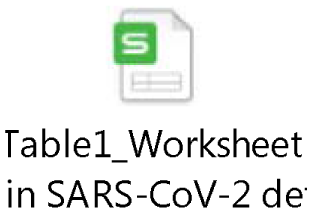
Data of RT-PCR, dPCR and CT results from 109 clinical specimens collected from 38 patients, including pharyngeal swab, stool and serum.

**Table 2.** RT-PCR and dPCR results of S1-S8 specimens

|  | RT-PCR (Ct) |  | dPCR result (copies/μl) |  |
| --- | --- | --- | --- | --- |
|  | ORF1ab gene | N gene | ORF1ab gene | N gene |
| S1 | 35 | 33 | 5.24 | 9.54 |
| S2 | 38 | 35 | 0.88 | 1.57 |
| S3 | undetermined | 38 | 0.10 | 0.10 |
| S4 | undetermined | undetermined | 0.10 | 0.10 |
| S5 | undetermined | undetermined | 0 | 0 |
| S6 | undetermined | undetermined | 0 | 0 |
| S7 | undetermined | undetermined | 0 | 0 |
| S8 | undetermined | undetermined | 0 | 0 |

### 3.2. Comparison of dPCR assay with RT-PCR assay using clinical samples

A total of 108 samples were taken from 38 individuals (**Table *1***) at various time points during the course of their treatment and quarantine. When assessed with RT-PCR method, the result showed reasonable consistency between different targets (**Table 3**).

**Table 3.**
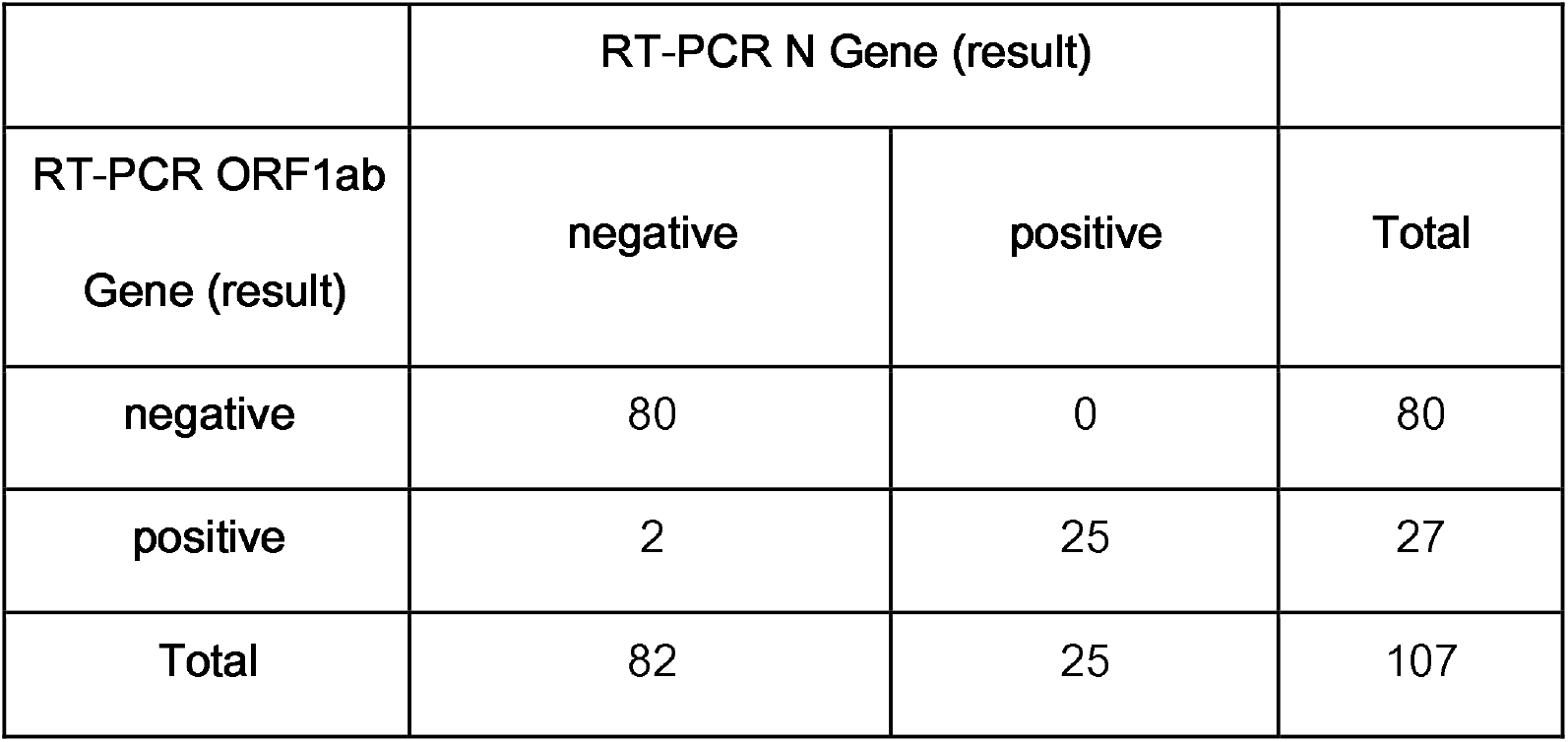
Consistency between RT-PCR assays targeting ORF1ab gene and N gene.

Results from dPCR targeting ORF1ab gene were also consistent with that from RT-PCR result (**Table *4***). In particular, 4 positive samples from 3 patients based on dPCR assay were not detected by RT-PCR. The computerized tomography (CT) results of the 3 patients showed 1 patient’s lung texture thickening, no pneumonia imaging characteristics; 1 patient had two lungs infections,; and 1 patient showed inflammation of the right lung and lower lobe. But RT-PCR assay failed to detect these 4 samples, and generated negative results.

**Table 4.** Consistency between dPCR assay targeting ORF1ab gene and RT-PCR assay targeting the same.

|  | dPCR ORF1ab (result) |  |  |
| --- | --- | --- | --- |
| RT-PCR ORF1ab | Negative | Positive | Total |

| Gene (result) |  |  |  |
| --- | --- | --- | --- |
| negative | 76 | 4 | 80 |
| positive | 3 | 25 | 28 |
| Total | 79 | 29 | 108 |

Digital PCR targeting N gene exhibited higher sensitivity than that targeting ORF1ab gene. As shown in **Table *5***, among 47 positive samples as determined by N gene, only 30 were positive as determined by ORF1ab gene. For the remaining 17 samples, 15 of them were from patients clinically determined to be positive based on other factors. One was a suspected case. <u>One</u> of them were of unknown clinical classification. In particular, one was actually discharged based on RT-PCR assay result, although a retrospective dPCR test detected the virus. In contrast, as long as a sample is deemed positive by ORF1ab gene, all but one were positive by N gene.

**Table 5.** Sensitivity of dPCR assays targeting ORF1ab gene and N gene.

|  | dPCR N gene (result) |  |  |
| --- | --- | --- | --- |
| dPCR ORF1ab gene (result) | Negative | Positive | Total |
| Negative | 62 | 17 | 79 |
| Positive | 0 | 30 | 30 |
| Total | 62 | 47 | 109 |

### 3.3. dPCR assay result during the course of treatment

Digital PCR also provides a window to monitor the progression and treatment of disease more consistently than RT-PCR. We analyzed positive samples from 3 patients based on dPCR assay that were not consistently detected by RT-PCR.

Patient 33 was first identified as SAR-CoV-2 positive by RT-PCR from pharyngeal swab with ORF1ab gene Ct of 32.3 and N gene Ct of 33.2 (**Table *6***). dPCR showed ORF1ab concentration of 28.3 copies/µl, N gene concentration of 35.2 copies/µl and internal reference gene concentration of 32.8 copies/µl on January 27, 2020 (**Figure *2*** and **Table *6***). The patient was treated in hospital under quarantine. The RT-PCR results from the same patient’s pharyngeal swab specimen were undetermined for both ORF1ab and N gene on February 5, 2020, after being treated for 9 days. dPCR results showed ORF1ab gene concentration of 0.44 copies/µl, N gene concentration of 0.62 copies/µl and internal reference gene of 130.97 copies/ µl. The ratio between ORF1ab gene and internal references gene decreased from 0.86 to 0.003 and the ratio between N gene and internal references gene decreased from 1.07 to 0.005 from January 27 to February 5. The dPCR results showed significant but incomplete viral clearance. Chest computed tomography (CT) results of the patient on January 27 and February 5 were both normal, showing no infection. The patient showed no fever, coughing, cold, muscle pain, pharynx, chest pain, diarrhea or nausea. dPCR was able to detect residual SARS-CoV-2 virus load from this asymptomatic patient.

**Table 6.**
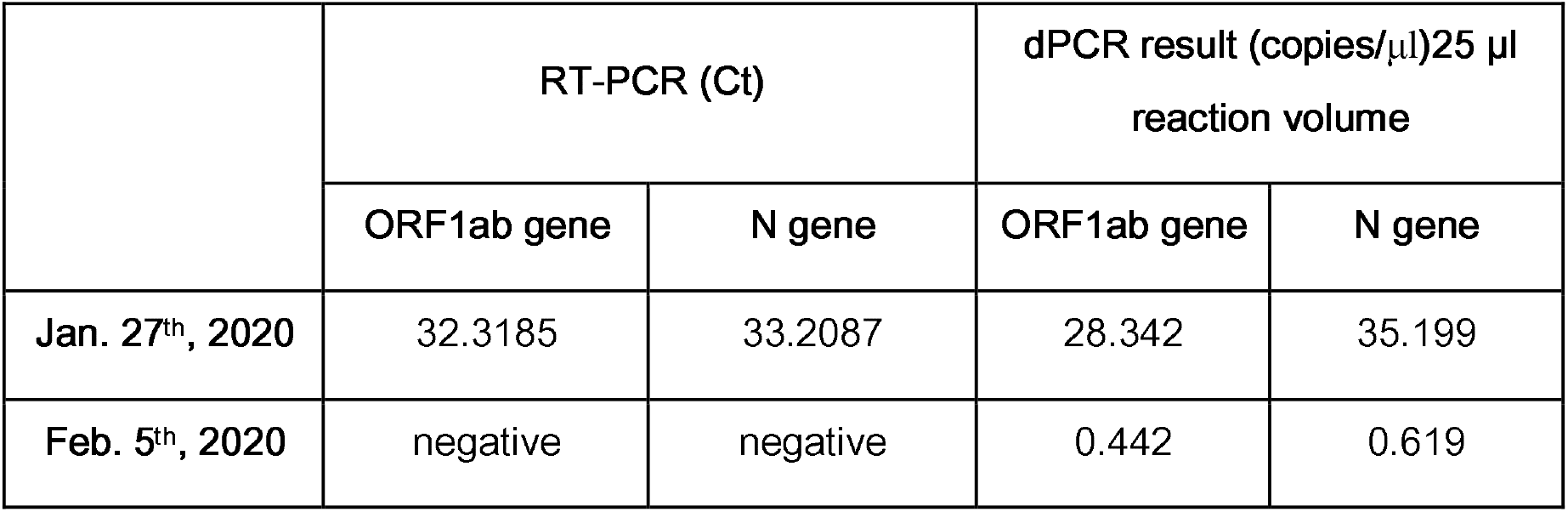
RT-PCR and dPCR results of Patient 33 on Jan. 27^th^, 2020 and Feb. 5^th^, 2020.

RT-PCR and dPCR results from Patient 8 on Jan. 23, 2020 both showed high SARS-CoV-2 viral load from the pharyngeal swab (**Table *7*** and **Figure 3**). The patient’s specimens were subsequently tested by RT-PCR and dPCR on February 5th and February 21. After 13 days of treatment (Feb. 5^th^), RT-PCR couldn’t detect any virus from pharyngeal swab. However, follow-up tests performed after another 16 days (Feb. 21^st^), RT-PCR results turned out to be positive from pharyngeal swab, but negative on stool specimen. CT results of this patient showed lung texture thickening without features characteristic of pneumonia. The patient also showed symptoms of fever, coughing, muscle pain and headache. The inconsistent results would confuse clinicians for diagnosis and treatment plan. dPCR assay, however, consistently reported positive results over the whole course of 29 days for specimens from pharyngeal swab. The ratio of ORF1ab gene and internal reference gene from pharyngeal swab by dPCR from patient 8 decreased from 1.66 to 0.001 from Jan. 23^rd^ to Feb. 21^st^. The ratio of N gene and internal reference gene from pharyngeal swab by dPCR from patient 8 decreased from 2.41 to 0.0007 from Jan. 23^rd^ to Feb. 21^st^. RT-PCR failed to detected the trace amount of SARS-CoV-2 virus from stool on Feb. 21^st^ (ORF1ab gene Ct > 41, N gene Ct undetermined). dPCR results were positive for patient 8’s stool specimen with ORF1ab gene concentration of 0.72 copies/µl and N gene concentration of 0.93 copies/µl.

**Table 7.** RT-PCR and dPCR results of patient 8 on Jan. 23^rd^, 2020, Feb. 5^th^, 2020 and Feb. 21^st^, 2020.

|  | RT-PCR (Ct) |  | dPCR (copies/μl)25 μl reaction volume |  | Ratio of genes from dPCR assay results |  |
| --- | --- | --- | --- | --- | --- | --- |
|  | ORF1ab gene | N gene | ORF1ab gene | N gene | ORF1ab/Reference | N/Reference |
| **Jan. 23rd, 2020** | 22.0 | 33.0 | 301.0 | 436.4 | 1.66 | 2.41 |
| **Feb. 5th, 2020** | negative | negative | 0.18 | 0.26 | 0.0025 | 0.0022 |
| **Feb. 21st, 2020** | 34.6 | 39.0 | 0.29 | 0.19 | 0.0012 | 0.0008 |

**Figure 3.**
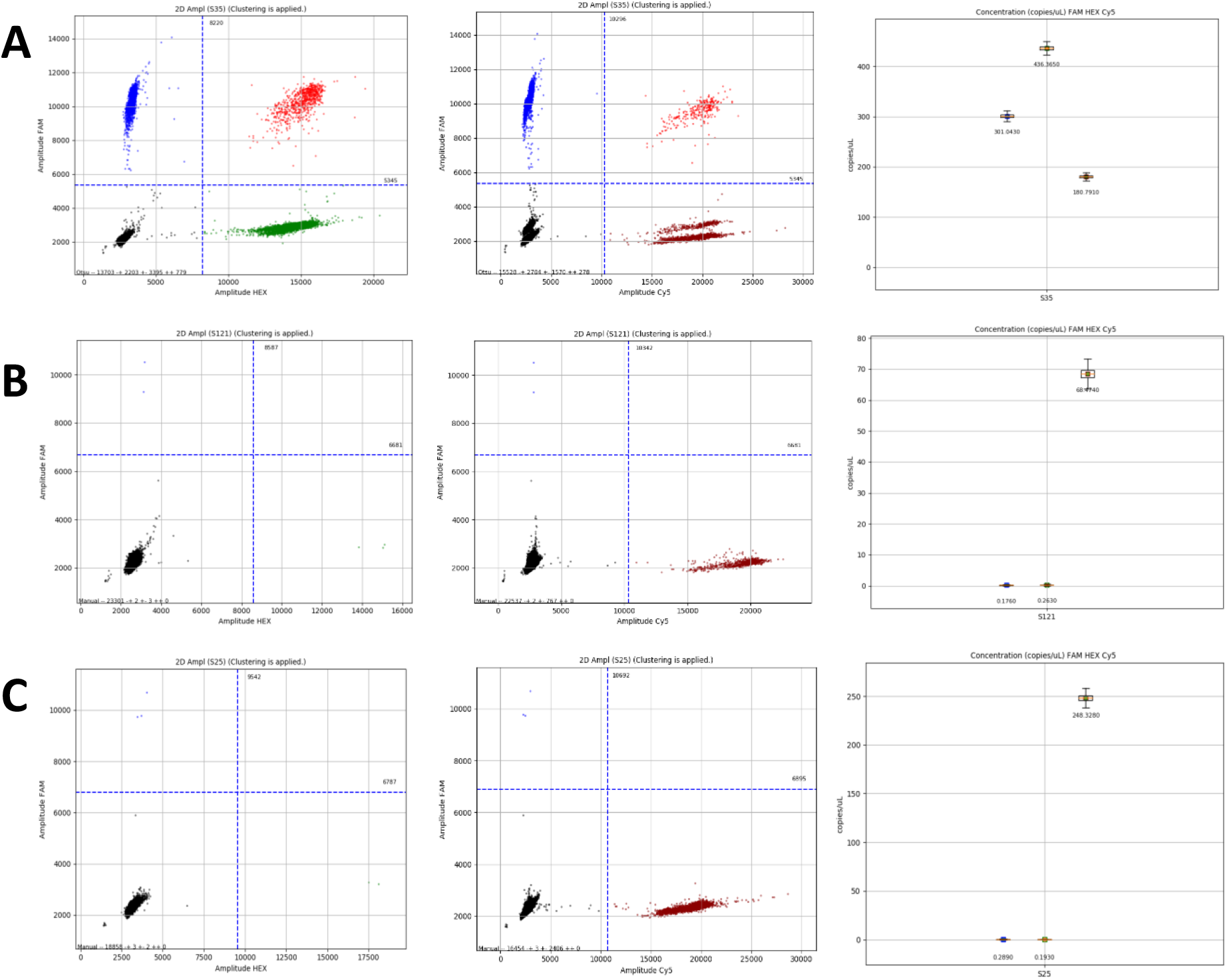
dPCR results of SAR-CoV-2 tests from patients 2. 3A is test result from Patient 8 on Jan. 23^rd^, 2020. 3B is test results from patient 8 on Feb. 5^th^, 2020. 3C is test results from patient 8 on Feb. 21^st^, 2020. Left is FAM (ORF1ab) Y axis, HEX (N) X axis; middle is FAM (ORF1ab) Y axis, Cy5 (internal reference) X axis; right is the concentration call of ORF1ab, N and internal reference genes.

Patient 9 first showed symptoms of fever, coughing and muscle pains on Feb. 4^th^, 2020 and then hospitalized on Feb. 13^th^, 2020. However, pharyngeal swabs were negative by RT-PCR from Feb. 14^th^ to Feb. 25^th^ for all 6 RT-PCR tests performed. CT results showed infection of both lungs.

However, due to the negative RT-PCR results, the patient was categorized as a suspected case despite her positive CT results and symptoms. Although dPCR showed negative result on Feb. 14^th^, the day after the patient intake, low SARS-CoV-2 viral load (ORF1ab gene concentration of 0.34 copies/µl and N gene of 0.43 copies/µl) was reported for the sample from Feb. 18^th^. The negative results of February 14^th^ were most likely due to insufficient sample loading. The concentration of internal reference gene was only 1.892 copies/μl, indicating low RNA load in the test. One Feb. 18^th^, dPCR test result was positive with the internal reference gene concentration at 20.8 copies/μl. It also suggested that the Feb. 14^th^ dPCR results were negative due to insufficient RNA loading which may be related to improper sample collection or RNA loss during sample nucleic acid extraction. CT scan result supported the conclusion from dPCR that this patient was infected with SARS-CoV-2. This patient was treated with Moxifloxacin, Arbidol and Pudilan.

These examples support our conclusion that dPCR offers improved sensitivity and consistency when testing specimens from patients during the course of treatment. dPCR is also able to detect low viral load in asymptomatic infection patients and suspected patients. dPCR can check if the negative result was caused by insufficient RNA loading by quantify the copy number of internal reference gene RPP30.

### 3.4. dPCR assay results from different sample locations

For 8 patients, both pharyngeal and stool samples were collected on the same day and tested. RT-PCR assay targeting ORF1ab gene reported positive result for all pharyngeal samples, but only 3 positive results for stool samples. dPCR assay targeting ORF1ab gene reported 7 positive results for pharyngeal samples, but only 1 positive result for stool samples. dPCR assay targeting N gene reported 8 positive results for pharyngeal samples, and 7 positive results for stool samples. Among these 8 patients, serum samples from 6 of them were also tested. The results were all negative when using RT-PCR and dPCR targeting ORF1ab genes, but dPCR targeting N gene reported 2 positive results. From dPCR results, it clearly indicates viral load in patients is throat > stool > blood.

The samples with ORF1ab positive detected in throat for dPCR is 6 more than RT-PCR (27 vs 21). The samples with ORF 1ab positive detected in blood and sputum for dPCR is 1 more than RT-PCR (1 vs 0). While the samples with ORF1ab positive detected in stool for dPCR is 2 less than RT-PCR (2 vs 4). Therefore, dPCR showed less sensitivity than RT-PCR in specimens from stool, higher sensitivity than RT-PCR in specimens from throat, blood and sputum. Different detection sensitivity of dPCR assay for different sample types may require further experiments to draw a conclusion.

### 3.5. dPCR assay results for internal reference gene

Stool specimen from patient 4 on Feb. 24^th^ was tested as negative by both RT-PCR and dPCR. However, the internal reference gene result by dPCR is 0 suggesting no RNA was loaded in either tests or the PCR reactions were inhibited. The samples need to be re-collected to re-run the tests. The internal reference gene of dPCR assay serves as a quality control to ensures no PCR inhibition happened and RNA extraction was successful. Without such a control in RT-PCR assay, the patient could have been discharged prematurely, putting those in close contact at risk of being infected.

## 4. Discussion

We first demonstrated that the LOD of dPCR assay is at least 10 times better than that of RT-PCR assay using serial dilution of the same clinical sample. Higher sensitivity, along with more reliable quantification of viral load, provides valuable information to help clinicians choose the appropriate treatment plan^16^. To demonstrate its application in clinical settings, we performed head-to-head comparison of RT-PCR and dPCR assays using a cohort of 39 patients totaling 109 samples obtained at different stage of the treatment and from different locations..

We observed that the result of dPCR targeting ORF1ab gene is consistent with RT-PCR targeting the same. In particular, dPCR assay detected 4 positive samples that were determined to be negative by RT-PCR assay. Although there were also 3 positive samples from RT-PCR assay that were deemed negative by dPCR-ORF1ab assay (the Ct values of ORF1ab for those 3 positive samples were > 38, which are in the grey area of RT-PCR), they were all positive from dPCR assay targeting N gene. Combing the CT results and test history of the 3 positive samples, 1 of the positive samples are false positive by RT-PCR (Patient 7).Combining the result of both dPCR assays provides a more sensitivity and accurate method to detect SARS-COV-2.

The dPCR results also showed higher sensitivity when the primer and probe are designed against N gene. It may be explained by the higher copy number of RNA for N gene arising from the replication process of the virus^17,18^. Interestingly, RT-PCR assay didn’t exhibit significance difference between the two targets. Further study is warranted to elucidate the underlying mechanism. It is well recognized that dPCR assays are less susceptible to the existence of PCR inhibitors, and the results are thus more reliable in general^11,19^.

With dPCR assay, we were able to track the progress of the treatment by monitoring the viral load from samples obtained on different dates. RT-PCR suffered from sporadic appearance of positive result which puzzled clinicians. dPCR results, in contrast, faithfully reflected the onset and healing of the disease, when examined together with relevant radiological evidence and treatment history. dPCR showed evidences of higher sensitivity to detect low virus load in patients who showed mild symptoms or have been treated for COVID-19 than RT-PCR.

We were also able to compare the viral load in different organs thanks to higher sensitivity of dPCR assay. The dPCR assay provides quantitative information on the viral load of specimens collected from different locations of the same patient. In all but one cases, the viral load is the highest in pharyngeal samples, lower in stool samples and the lowest in serum. Interestingly, a considerable amount of virus was found in the phlegm of one of the patients whose pharyngeal sample was negative. These observations may provide valuable insight into the pathology of this emerging disease^20^.

## 5. Conclusions

Digital PCR shows improved lower limit of detection, sensitivity and accuracy, enabling COVID-19 detection with less false negative and false positive results comparing with RT-PCR, especially for the tests with low viral load specimens. We showed evidences that dPCR is powerful in detecting asymptomatic patients and suspected patients. Digital PCR is capable of flagging the negative results caused by insufficient sample loading by quantifying internal reference gene from human RNA in the PCR reactions. Multi-channel fluorescence dPCR system (FAM/HEX/CY5/ROX) is able to detect more target genes in a single multiplex assay, providing quantitative count of viral load in specimens, which is a powerful tool for monitoring COVID-19 treatment.

## Data Availability

3/25/2020

## Author Contributions

Weihua Cai conceptualized the study design; Renfei Lu, Jian Wang recruited the patients, collected specimens, did the laboratory tests and interpreted the results; Jia Dong did the part of the laboratory tests. Yaqi Wang analyzed the data; Renfei Lu wrote the drafts of the manuscript; all authors read and approved the final report and draft.

## Acknowledgement

We are grateful to RainSure Scientific for their great support to this work. We are grateful to Drs. Yu Liu, Drs. Yunfeng Ling, Drs. Chen Li, Drs. Xuan Zhang and Mr. Haifeng Wang, Mr. Yajun Xu and Mr. Hua Zhang for their support in experiment design, data analysise and instrumentation.

